# Sleep-Disordered Breathing Profiles in Patients with Cardiovascular Diseases: Kurume SDB-CVD Study

**DOI:** 10.1101/2024.06.11.24308799

**Authors:** Koutatsu Shimozono, Hisashi Adachi, Shoichiro Nohara, Tatsuhiro Shibata, Yoichi Sugiyama, Tetsuya Ioji, Yoshihiro Fukumoto

**Author notes:** **Corresponding author** Yoshihiro Fukumoto, MD, PhD, FJCS, FJCC, FAHA, FESC., Division of Cardiovascular Medicine, Department of Internal Medicine, Kurume University School of Medicine, 67 Asahi-machi, Kurume, 830-0011, Japan.

## Abstract

**Background:** While evidence links sleep-disordered breathing (SDB) to various cardiovascular diseases (CVDs), comprehensive insights and detailed profiles of CVD patients with SDB remain scarce. This study aimed to examine the characteristics of CVD patients complicated by SDB.

**Methods:** This retrospective study enrolled all consecutive patients who underwent screening tests for SDB during hospitalization at Kurume University Hospital from January 2014 to December 2019. To explore the relationship between SDB severity and clinical data, we classified SDB severity into four groups and assessed its prevalence and severity across different CVDs, categorized by sex and age.

**Results:** A total of 5,765 patients were enrolled. Desaturation during sleep in SDB was significantly worse in terms of age, sex, daytime resting oxygen saturation (SpO_2_), systolic blood pressure (SBP), pulse pressure, body mass index (BMI), white blood cell count, hemoglobin A1c, serum creatinine, N-terminal Pro-B-type natriuretic peptide (NT-proBNP), cardiothoracic ratio on chest X-ray, and left ventricular ejection fraction (LVEF) on echocardiography (all p-values < 0.001, except for SBP p=0.002, and NT-proBNP p=0.004). Daytime SpO_2_ and BMI strongly correlated across ages. Differences in renal function, diabetes control, and LVEF were notable between sex and ages. Analysis also showed the more prevalence and severity of SDB in coronary artery disease and heart failure, versus lower in pulmonary hypertension. Heatmaps of daytime SpO2 and BMI can effectively predict the severe SDB in patients with CVDs, suggesting the potential for efficient therapeutic interventions for SDB.

**Conclusion:** Coronary artery disease and heart failure were associated with a higher prevalence and a greater risk of moderate to severe SDB, typically in older patients. Notably, low SpO_2_ during daytime and elevated BMI were indicative of SDB across all age categories for both sexes.

## Introduction

Sleep-disordered breathing (SDB) is a condition characterized by abnormal respiratory patterns during sleep, encompassing entities primarily defined by obstructive phenomena -such as repetitive narrowing and closure of the upper airway- as well as disorders marked by neuromuscular output irregularities without significant airway obstruction.^1^ SDB is linked to a cascade of pathophysiological outcomes including intermittent hypoxia, oxidative stress, sympathetic nervous system activation, and endothelial dysfunction, contributing to the development of hypertension, coronary artery disease, arrhythmias, heart failure, and stroke.^2^ While normal sleep reduces physiological stress, beneficially impacting the cardiovascular system, SDB disrupts these restorative sleep processes. Both obstructive and central sleep apneas exhibit apneas, hypopneas, and subsequent compensatory hyperpneas, leading to arterial blood gas perturbations, frequent arousals, a shift towards sympathetic dominance, and significant negative intrathoracic pressure fluctuations.^2^

SDB prevalence is high, with moderate-to-severe obstructive sleep apnea (OSA) affecting an estimated 20% of adult males and 10% of postmenopausal females globally, extending beyond Western populations to include countries such as Japan.^3^ The worldwide prevalence of OSA affects approximately 1 billion individuals, with rates surpassing 50% in some countries.^4^ In Japan, an estimated 9.4 million people exhibit an apnea-hypopnea index (AHI) of ≥15 events per hour, representing 14.0% of the population aged 30-69 years.^3^ The adoption of continuous positive airway pressure (CPAP) therapy for OSA is rising sharply, nearing 500,000 users in Japan alone.^3^

Sleep apnea prevalence is notably higher among older individuals, males, and those with cardiovascular diseases (CVDs) and lifestyle-related conditions.^2,5^ However, comprehensive comparisons of SDB prevalence across different CVDs within a specific cohort are lacking. In our clinical practice, we routinely assess for SDB in patients with CVDs admitted to our institution using pulse oximetry. Consequently, this study aimed to investigate the prevalence of SDB among hospitalized patients for CVDs, stratified by age and sex, and to delineate the association between SDB and clinical profiles in each CVD category.

## Methods

### Study Design

This study was a retrospective cohort study using the database of the Department of Cardiovascular Medicine, Kurume University Hospital. We enrolled all consecutive patients, who have taken screening test of SDB by PULSOX®-Me300 (Konica Minolta, Inc., Japan) during hospitalization from January 2014 to December 2019. This oxygen saturation analyzer, PULSOX®-Me300, accurately measures transcutaneous arterial blood oxygen saturation (SpO_2_) by updating the data every second following the initiation of measurement.

The present study received ethical approval from the Ethics Committee of Kurume University Hospital (approval no. 20181). The informed consent was waived due to the retrospective nature and opt-out was used in the study. The study was conducted following the ethical principles outlined in the Declaration of Helsinki.

### Study population and protocol

SDB was evaluated using PULSOX®-Me300 in patients, excluding those admitted urgently due to acute illness or those unable to provide consent.

Of the 8628 patients enrolled in the study, 5765 were evaluable for 3% Oxygen Desaturation Index (ODI) using PULSOX®-Me300. They ranged in age from 13 to 100 years (mean 66.5 ± 14.4 years), with 3523 (61.1%) males and 2242 (38.9%) females. Patients were classified no SDB (3% ODI<5), mild SDB (5≤3% ODI<15), moderate SDB (15≤3% ODI<30), and severe SDB (30≤3% ODI).^6^

Considering the well-documented prevalence of SDB in older populations, we stratified patients into age groups of under 60, 60s, 70s, and 80s, and conducted subsequent analyses by sex within each age category. We assessed the prevalence and severity of SDB across various CVDs to identify those most significantly impacted by SDB.

### Data collection

Data were collected from the database, which consists of baseline demographic data such as age, sex, height, body weight, SpO_2_ in the awake state, systolic blood pressure, diastolic blood pressure, and pulse rate. Body mass index (BMI) was calculated as weight (kilograms) divided by the square of height (square meters) as an index of obesity.

Blood samples were obtained from the antecubital vein and measured at a commercially available laboratory in Kurume University Hospital [white blood cell count, hemoglobin, hemoglobin A1c, serum creatinine, serum sodium, N-terminal pro-B-type natriuretic peptide (NT-proBNP)]. Also, cardiothoracic ratio on plain chest x-ray and left ventricular ejection fraction (LVEF) measured using the 2D modified Simpson’s method were obtained.

We, 4 expert cardiologists, classified CVDs as coronary artery disease, arrhythmia, heart failure, valvular heart disease, pulmonary hypertensive heart failure, and others. Coronary artery disease included chronic stable angina, asymptomatic myocardial ischemia, acute coronary syndrome, previous myocardial infarction, previous coronary revascularization, coronary spastic angina, and nonocclusive coronary atherosclerosis. Arrhythmia was the primary clinical diagnosis, with patients admitted for atrial and/or ventricular arrhythmias, including atrial fibrillation and ventricular tachycardia. Heart failure is a complex clinical syndrome resulting from structural and functional abnormalities of the heart. The clinical diagnosis of heart failure was defined as a patient presenting with heart failure symptoms due to some cardiac disease. Valvular heart disease was diagnosed as the primary clinical diagnosis in patients admitted due to abnormalities of the aortic, mitral, tricuspid, or pulmonary valves (e.g., aortic stenosis, aortic regurgitation, mitral regurgitation). Pulmonary hypertension was diagnosed when the mean pulmonary arterial pressure at rest increased by 25 mmHg or more due to pulmonary arterial disease and chronic thromboembolic disease; pulmonary arterial hypertension and chronic thromboembolic pulmonary hypertension were also diagnosed. Diseases other than these five categories were classified as others.

### Heatmap

The heatmap illustrates the prevalence (%) of severe SDB patients and the relationship between BMI and SpO_2_. BMI data is divided into deciles, and SpO_2_ is segmented into ranges from 90 to 97 with increments of 1. The shading on the heatmap uses red to indicate the highest occurrence frequency and green to denote the lowest occurrence frequency.

### Statistical Analysis

Data were presented mean ± standard deviation (SD). Chi-square (χ2) tests were utilized to analyze categorical parameters and assess differences between groups. The mean values of the parameters, stratified by the SDB severity, were compared using analysis of variance (ANOVA). Univariate regression analysis, employing linear regression, was utilized to explore the associations between variables. Factors deemed significant in the univariate analysis were further evaluated through multiple regression analysis. This method enabled the sequential addition or removal of predictors based on their statistical significance, allowing for the determination of their independent contributions to the dependent variable. Variableness that maintained statistical significance in the ultimate model were recognized as having an independent association with the dependent variable. The Mantel-Haenszel test was employed to analyze the SDB severity across various CVDs. Significant differences in SDB severity were observed among types of CVDs, stratified by the entire cohort and by sex. P-values <0.05 were considered statistically significant. All statistical analyses were performed using SPSS version 23.0 (IBM Inc., Chicago, IL, USA).

## Results

### Characterization of Sleep-Disordered Breathing in Patients with Cardiovascular Diseases

A total of consecutive 8628 patients were enrolled in this study. Of them, 2,863 were excluded due to the lack of available PULSOX®-Me300 data, often resulting from emergency or acute care hospitalizations, and 5,765 patients were included. The age distribution of the included patients ranged from 13 to 100 years, with a mean age of 66.5 ± 14.4 years. The cohort comprised 3,523 (61.1%) males and 2,242 (38.9%) females (**Table 1**). Notably, significant sex differences were observed across all profiles except for pulse pressure and NT-proBNP levels, underscoring the importance of sex-specific evaluations to elucidate the characteristics of SDB.

**Table 1.**
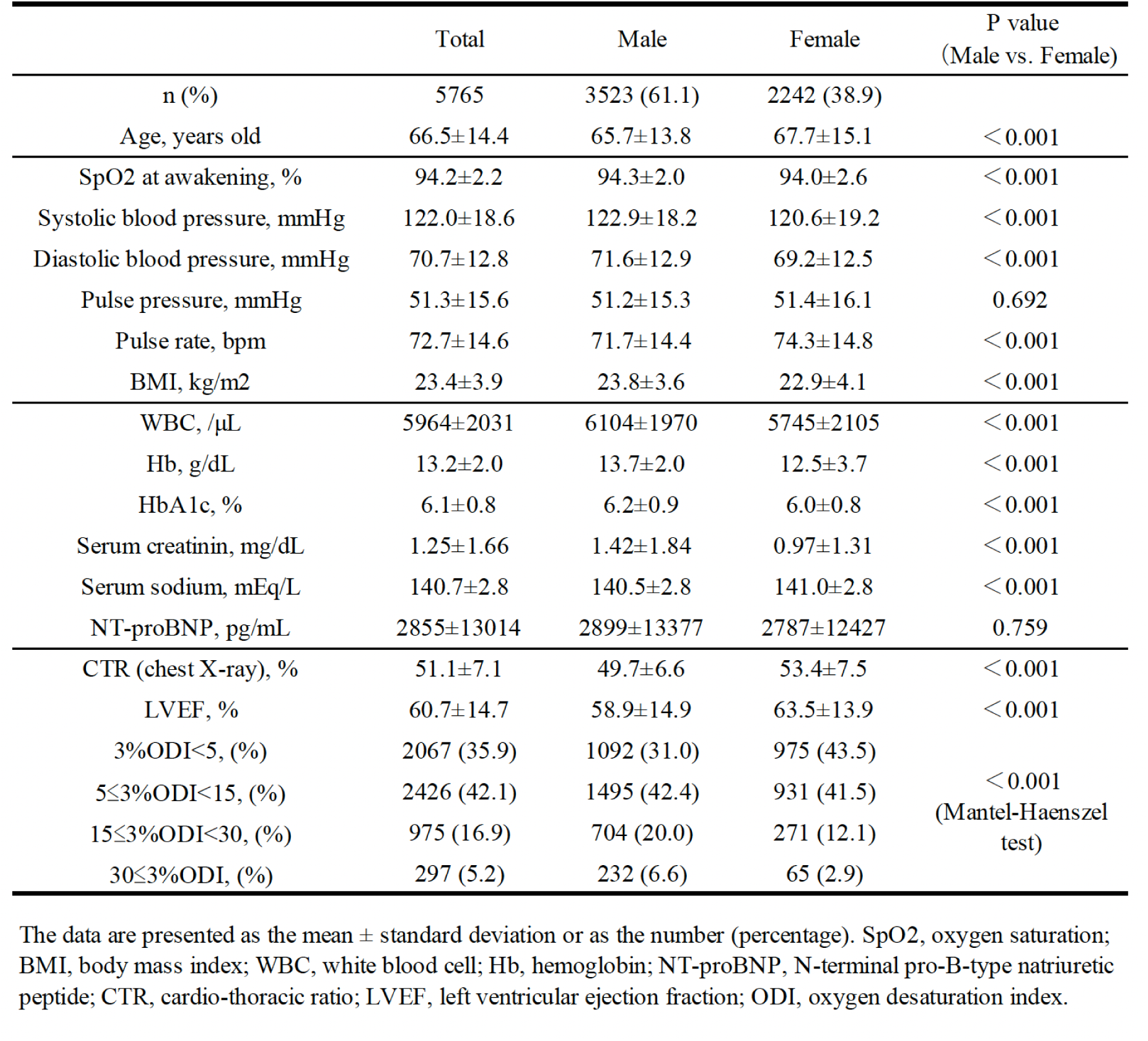
Baseline characteristics of all enrolled patients with cardiovascular diseases.

|  | Total | Male | Female | P value<br>(Male vs. Female) |
| --- | --- | --- | --- | --- |
| n (%) | 5765 | 3523 (61.1) | 2242 (38.9) |  |
| Age, years old | 66.5±14.4 | 65.7±13.8 | 67.7±15.1 | < 0.001 |
| SpO2 at awakening, % | 94.2±2.2 | 94.3±2.0 | 94.0±2.6 | < 0.001 |
| Systolic blood pressure, mmHg | 122.0±18.6 | 122.9±18.2 | 120.6±19.2 | < 0.001 |
| Diastolic blood pressure, mmHg | 70.7±12.8 | 71.6±12.9 | 69.2±12.5 | < 0.001 |
| Pulse pressure, mmHg | 51.3±15.6 | 51.2±15.3 | 51.4±16.1 | 0.692 |
| Pulse rate, bpm | 72.7±14.6 | 71.7±14.4 | 74.3±14.8 | < 0.001 |
| BMI, kg/m2 | 23.4±3.9 | 23.8±3.6 | 22.9±4.1 | < 0.001 |
| WBC, /μL | 5964±2031 | 6104±1970 | 5745±2105 | < 0.001 |
| Hb, g/dL | 13.2±2.0 | 13.7±2.0 | 12.5±3.7 | < 0.001 |
| HbA1c, % | 6.1±0.8 | 6.2±0.9 | 6.0±0.8 | < 0.001 |
| Serum creatinin, mg/dL | 1.25±1.66 | 1.42±1.84 | 0.97±1.31 | < 0.001 |
| Serum sodium, mEq/L | 140.7±2.8 | 140.5±2.8 | 141.0±2.8 | < 0.001 |
| NT-proBNP, pg/mL | 2855±13014 | 2899±13377 | 2787±12427 | 0.759 |
| CTR (chest X-ray), % | 51.1±7.1 | 49.7±6.6 | 53.4±7.5 | < 0.001 |
| LVEF, % | 60.7±14.7 | 58.9±14.9 | 63.5±13.9 | < 0.001 |
| 3%ODI<5, (%) | 2067 (35.9) | 1092 (31.0) | 975 (43.5) | < 0.001<br>(Mantel-Haenszel<br>test) |
| 5≤3%ODI<15, (%) | 2426 (42.1) | 1495 (42.4) | 931 (41.5) |  |
| 15≤3%ODI<30, (%) | 975 (16.9) | 704 (20.0) | 271 (12.1) |  |
| 30≤3%ODI, (%) | 297 (5.2) | 232 (6.6) | 65 (2.9) |  |

The data are presented as the mean ± standard deviation or as the number (percentage). SpO2, oxygen saturation; BMI, body mass index; WBC, white blood cell; Hb, hemoglobin; NT-proBNP, N-terminal pro-B-type natriuretic peptide; CTR, cardio-thoracic ratio; LVEF, left ventricular ejection fraction; ODI, oxygen desaturation index.

Patients were categorized into 4 groups based on SDB severity: no SDB, mild SDB, moderate SDB, and severe SDB. Consistent with prior findings, more severe SDB was predominantly observed in older males (**Tables 1 and 2**). Remarkably, SpO_2_ upon awakening demonstrated a significant decline with increasing SDB severity (no SDB; 95.1±1.9%, mild SDB; 94.0±2.0%, moderate SDB; 93.2±2.1%, and severe SDB; 91.9±3.3%, P<0.001, **Table 2**). Furthermore, patients with more severe SDB exhibited higher pulse pressure, increased BMI, elevated white blood cell counts, reduced hemoglobin levels, higher HbA1c, elevated serum creatinine and NT-proBNP levels, greater instances of cardiomegaly, and decreased LVEF (**Table 2**).

**Table 2.** Baseline characteristics by 3% ODI.

| All | 3%ODI<5 | 5≤3%ODI<15 | 15≤3%ODI<30 | 30≤3%ODI | P for trend |
| --- | --- | --- | --- | --- | --- |
| n (%) | 2067 (35.9) | 2426 (42.1) | 975 (16.9) | 297 (5.2) |  |
| Male/Female (Male,%) | 1092/975 (52.8) | 1495/931 (61.6) | 704/271 (72.2) | 232/65 (78.1) | <0.001 |
| Age, years old | 62.4±16.8 | 67.6±12.7 | 70.8±11.2 | 70.9±11.5 | <0.001 |
| SpO2 at awakening, % | 95.1±1.9 | 94.0±2.0 | 93.2±2.1 | 91.9±3.3 | <0.001 |
| Systolic blood pressure, mmHg | 121.5±18.7 | 121.7±18.8 | 122.6±18.0 | 125.6±18.5 | 0.002 |
| Diastolic blood pressure, mmHg | 70.9±12.6 | 70.4±12.8 | 70.4±12.8 | 72.5±13.1 | 0.589 |
| Pulse pressure, mmHg | 50.6±15.5 | 51.3±15.5 | 52.2±15.9 | 53.1±16.8 | 0.001 |
| Pulse rate, bpm | 72.9±14.4 | 72.6±14.3 | 72.2±15.3 | 74.4±15.4 | 0.871 |
| BMI, kg/m2 | 22.3±3.4 | 23.6±3.6 | 24.6±4.0 | 25.9±5.1 | <0.001 |
| WBC, /μL | 5791±1876 | 6013±2189 | 6092±1924 | 6341±1979 | <0.001 |
| Hb, g/dL | 13.1±1.9 | 13.2±2.0 | 13.1±2.1 | 13.3±2.2 | 0.222 |
| HbA1c, % | 6.0±0.8 | 6.1±0.8 | 6.3±0.9 | 6.4±0.9 | <0.001 |
| Serum creatinin, mg/dL | 1.13±1.54 | 1.26±1.74 | 1.36±1.66 | 1.56±1.88 | <0.001 |
| Serum sodium, mEq/L | 140.6±2.9 | 140.8±2.8 | 140.6±3.0 | 140.8±2.6 | 0.767 |
| NT-proBNP, pg/mL | 2551±11155 | 2674±13191 | 3148±13353 | 5533±202 | 0.004 |
| CTR (chest X-ray), % | 49.9±7.6 | 51.3±6.9 | 52.3±6.8 | 53.8±6.2 | <0.001 |
| LVEF, % | 62.4±13.5 | 60.7±14.9 | 58.8±15.4 | 55.2±16.3 | <0.001 |
| Male | 3%ODI<5 | 5≤3%ODI<15 | 15≤3%ODI<30 | 30≤3%ODI | P for trend |
| n (%) | 1092 (31.0) | 1495 (42.4) | 704 (20.0) | 232 (6.6) |  |
| Age, year | 61.5±16.5 | 66.2±12.4 | 69.7±10.7 | 69.7±11.3 | <0.001 |
| SpO2 at awakening, % | 95.2±1.6 | 94.3±1.8 | 93.4±2.0 | 92.2±2.3 | <0.001 |
| Systolic blood pressure, mmHg | 122.9±18.1 | 122.5±18.5 | 122.9±18.0 | 124.7±18.2 | 0.349 |
| Diastolic blood pressure, mmHg | 72.3±12.7 | 71.3±12.8 | 71.0±13.2 | 72.4±12.7 | 0.26 |
| Pulse pressure, mmHg | 50.6±15.1 | 51.2±15.2 | 51.9±15.5 | 52.3±16.0 | 0.039 |
| Pulse rate, bpm | 71.6±14.4 | 71.6±14.1 | 71.2±14.5 | 74.5±15.5 | 0.174 |
| BMI, kg/m2 | 22.7±3.1 | 23.8±3.4 | 24.8±3.8 | 26.1±4.7 | <0.001 |
| WBC, /μL | 5909±1767 | 6164±2116 | 6181±1931 | 6399±1952 | <0.001 |
| Hb, g/dL | 13.8±1.9 | 13.7±2.0 | 13.5±2.1 | 13.6±2.1 | 0.003 |
| HbA1c, % | 6.0±0.8 | 6.2±0.8 | 6.3±0.9 | 6.5±0.8 | <0.001 |
| Serum creatinin, mg/dL | 1.31±1.78 | 1.46±1.88 | 1.46±1.80 | 1.63±1.93 | 0.011 |
| Serum sodium, mEq/L | 140.5±2.9 | 140.6±2.8 | 140.5±3.0 | 140.7±2.5 | 0.468 |
| NT-proBNP, pg/mL | 2628±12001 | 2790±13934 | 2670±10224 | 5582±21563 | 0.057 |
| CTR (chest X-ray), % | 47.7±6.5 | 49.8±6.4 | 51.4±6.5 | 52.8±5.7 | <0.001 |
| LVEF, % | 61.2±13.4 | 58.8±15.2 | 57.1±15.4 | 53.4±16.2 | <0.001 |
| Female | 3%ODI<5 | 5≤3%ODI<15 | 15≤3%ODI<30 | 30≤3%ODI | P for trend |
| n (%) | 975 (43.9) | 918 (41.3) | 266 (12.0) | 62 (2.8) |  |
| Age, year | 63.4±17.0 | 69.9±12.9 | 73.9±11.0 | 75.4±10.9 | <0.001 |
| SpO2 at awakening, % | 94.9±2.2 | 93.6±2.3 | 92.7±2.3 | 90.8±5.5 | <0.001 |
| Systolic blood pressure, mmHg | 120.0±19.4 | 120.3±19.3 | 121.8±17.9 | 128.7±19.2 | 0.005 |
| Diastolic blood pressure, mmHg | 69.3±12.4 | 68.9±12.7 | 68.8±11.3 | 73.0±14.3 | 0.555 |
| Pulse pressure, mmHg | 50.7±15.9 | 51.4±15.9 | 53.0±16.8 | 55.8±19.4 | 0.004 |
| Pulse rate, bpm | 74.3±14.3 | 74.2±14.5 | 74.9±17.0 | 74.0±15.5 | 0.827 |
| BMI, kg/m2 | 21.9±3.8 | 23.3±3.9 | 24.2±4.5 | 25.2±6.4 | <0.001 |
| WBC, /μL | 5660±1983 | 5773±2280 | 5865±1892 | 6133±2105 | 0.033 |
| Hb, g/dL | 12.5±1.7 | 12.4±1.7 | 12.3±1.7 | 12.3±2.2 | 0.64 |
| HbA1c, % | 5.9±0.8 | 6.0±0.7 | 6.3±0.7 | 6.3±1.0 | <0.001 |
| Serum creatinin, mg/dL | 0.93±1.17 | 0.96±1.43 | 1.09±1.20 | 1.30±1.67 | 0.016 |
| Serum sodium, mEq/L | 140.9±2.8 | 141.1±2.7 | 141.0±3.0 | 141.1±3.0 | 0.227 |
| NT-proBNP, pg/mL | 2465±10143 | 2484±11889 | 4387±19210 | 5358±15124 | 0.023 |
| CTR (chest X-ray), % | 52.3±7.9 | 53.8±7.1 | 54.7±6.8 | 57.3±6.5 | <0.001 |
| LVEF, % | 63.6±13.7 | 63.6±14.0 | 63.3±14.6 | 61.8±15.0 | 0.469 |

Subsequent analyses focused on sex-differences revealed that both males and females with more severe SDB were older, had lower SpO_2_ at awakening, higher pulse pressure, larger BMI, higher WBC counts, increased HbA1c, elevated serum creatinine and NT-proBNP levels, and more frequent cardiomegaly (**Table 2**). Among males, more severe SDB was significantly associated with reduced LVEF, but not in females (**Table 2**).

To further investigate age-related differences, the cohort was stratified by age groups -under 60, 60s, 70s, and over 80- as well as by sex, facilitating a more nuanced analysis of the interplay between age, sex, and SDB severity.

### Analysis of Sleep-Disordered Breathing Severity Across Age Groups in All Enrolled Patients

In examining SDB severity across various age groups, analyses were performed for combined sexes, males alone, and females alone, yielding consistent findings. Across these groups, more severe forms of SDB were invariably linked with lower SpO_2_ upon awakening and increased BMI (**Supplemental Tables 1-4**). Additionally, elevated HbA1c levels correlated with SDB severity in all demographics, except female at the age of 70 years or older. The association of cardiomegaly and decreased LVEF with SDB was more pronounced in males than in females (**Supplemental Tables 1-4**).

### Association Between Cardiovascular Diseases and Sleep-Disordered Breathing in the Enrolled Population

Within the studied cohort, the prevalence of CVDs was highest for coronary artery disease at 28.8%, followed by arrhythmias at 25.9%, heart failure at 22.1%, valvular heart disease at 6.3%, pulmonary hypertension at 5.8%, and other conditions at 11.1%. Among males, coronary artery disease prevalence increased to 34.7%, with subsequent prevalences for arrhythmias, heart failure, valvular heart disease, pulmonary hypertension, and other conditions aligning with the overall trends. Conversely, in females, arrhythmias were most common at 24.6%, followed by heart failure, coronary artery disease, pulmonary hypertension, valvular heart disease, and other conditions, displaying a unique distribution pattern (**Figure 1**).

**Figure 1.**
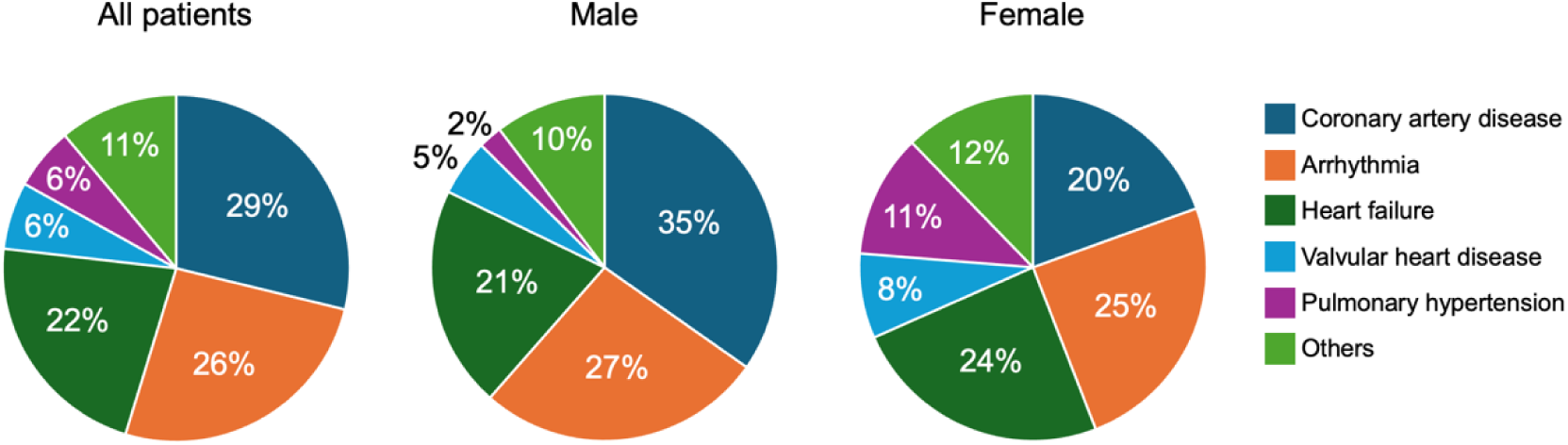
Underlying cardiovascular diseases in this study database. Left; the overall population, middle; male, right; female. The top 3 major diseases were coronary artery disease, arrhythmia, and heart failure.

Given the significant correlation between SDB severity and factors such as sex, age, SpO_2_ at awakening, BMI, and HbA1c levels, we explored the prevalence trends of underlying CVDs in relation to SDB. Valvular heart disease patients were generally older, the lowest SpO_2_ levels were observed in pulmonary hypertension, the highest BMI was noted in coronary artery disease, and elevated HbA1c levels were most prevalent among patients with coronary artery disease (**Table 3**).

**Table 3.** Baseline characteristics of each cardiovascular disease.

| All | Coronary artery disease | Arrhythmia | Heart failure | Valvular heart disease | Pulmonary hypertension | Others | P for trend |
| --- | --- | --- | --- | --- | --- | --- | --- |
| n (%) | 1661 (28.8) | 1494 (25.9) | 1273 (22.1) | 362 (6.3) | 335 (5.8) | 640 (11.1) |  |
| Male/Female (Male,%) | 1224/437 (73.7) | 942/552 (63.1) | 729/544 (57.3) | 185/177 (51.1) | 78/257 (23.3) | 365/275 (57.0) | <0.001 |
| Age, years old | 69.3±11.2 | 62.0±14.8 | 70.3±13.5 | 73.1±12.6 | 61.3±16.5 | 60.8±16.1 | <0.001 |
| SpO2 at awakening, % | 94.2±2.0 | 94.5±1.8 | 94.0±2.5 | 94.3±1.9 | 92.3±3.6 | 94.3±2.2 | <0.001 |
| Systolic blood pressure, mmHg | 124.3±17.6 | 123.7±16.3 | 118.4±20.8 | 125.4±19.2 | 111.5±17.3 | 122.6±19.2 | <0.001 |
| Diastolic blood pressure, mmHg | 69.6±11.5 | 74.5±12.1 | 67.9±13.8 | 70.0±12.9 | 66.8±11.7 | 72.4±13.4 | <0.001 |
| Pulse pressure, mmHg | 54.7±14.7 | 49.2±14.7 | 50.4±17.4 | 55.5±16.3 | 44.7±12.9 | 50.2±14.7 | <0.001 |
| Pulse rate, bpm | 69.6±11.9 | 71.8±16.5 | 74.8±15.2 | 73.3±12.1 | 81.2±13.7 | 74.0±14.0 | <0.001 |
| BMI, kg/m2 | 24.0±3.7 | 23.8±3.6 | 23.1±4.1 | 22.1±3.4 | 22.1±4.4 | 23.1±4.0 | <0.001 |
| WBC, /μL | 6234±1995 | 5666±1546 | 6144±2468 | 5732±1852 | 5388±2052 | 6026±2075 | <0.001 |
| Hb, g/dL | 13.1±1.8 | 14.0±1.7 | 12.6±2.2 | 12.4±1.8 | 12.4±2.0 | 13.3±1.9 | <0.001 |
| HbA1c, % | 6.4±1.0 | 5.9±0.6 | 6.2±0.9 | 5.9±0.7 | 5.9±0.7 | 5.9±0.6 | <0.001 |
| Serum creatinin, mg/dL | 1.48±1.99 | 0.93±1.22 | 1.47±1.79 | 1.30±1.75 | 0.85±0.79 | 1.11±1.48 | <0.001 |
| Serum sodium, mEq/L | 140.7±2.7 | 141.4±2.1 | 140.0±3.6 | 140.5±2.9 | 140.8±2.7 | 140.7±2.6 | <0.001 |
| NT-proBNP, pg/mL | 2472±12718 | 617±2000 | 6759±20604 | 3645±13486 | 867±1669 | 1792±9728 | <0.001 |
| CTR (chest X-ray), % | 49.1±5.7 | 48.3±5.9 | 56.1±7.5 | 52.9±6.6 | 52.8±7.2 | 51.1±7.3 | <0.001 |
| LVEF, % | 61.0±12.8 | 65.9±8.5 | 50.0±18.8 | 64.5±11.7 | 68.9±8.5 | 62.9±13.6 | <0.001 |
| Male | Coronary artery disease | Arrhythmia | Heart failure | Valvular heart disease | Pulmonary hypertension | Others | P for trend |
| n (%) | 1224 (34.7) | 942 (26.7) | 729 (20.7) | 185 (5.3) | 78 (2.2) | 365 (10.4) |  |
| Age, year | 68.8±10.8 | 60.7±14.6 | 68.3±13.6 | 69.4±12.5 | 64.1±17.4 | 61.4±15.8 | <0.001 |
| SpO2 at awakening, % | 94.3±1.8 | 94.5±1.7 | 94.1±2.4 | 94.5±2.0 | 92.3±3.6 | 94.3±1.8 | <0.001 |
| Systolic blood pressure, mmHg | 124.0±17.3 | 123.8±15.7 | 118.5±21.0 | 125.6±18.5 | 115.8±20.4 | 125.3±19.1 | <0.001 |
| Diastolic blood pressure, mmHg | 70.1±11.5 | 75.5±12.1 | 68.6±14.0 | 71.1±13.7 | 67.6±11.7 | 73.9±13.6 | <0.001 |
| Pulse pressure, mmHg | 53.9±14.3 | 48.3±14.1 | 49.9±17.2 | 54.4±15.9 | 48.2±15.6 | 51.4±14.8 | <0.001 |
| Pulse rate, bpm | 69.3±12.1 | 71.8±16.1 | 74.0±15.2 | 72.1±13.0 | 81.0±14.5 | 72.7±13.9 | <0.001 |
| BMI, kg/m2 | 24.2±3.4 | 24.2±3.5 | 23.5±3.9 | 22.2±3.2 | 21.6±3.7 | 23.5±3.8 | <0.001 |
| WBC, /μL | 6324±1987 | 5727±1546 | 6258±2282 | 5732±1968 | 5793±2134 | 6265±2006 | <0.001 |
| Hb, g/dL | 13.4±1.8 | 14.6±1.5 | 13.1±2.3 | 12.8±1.9 | 13.1±2.4 | 13.8±1.9 | <0.001 |
| HbA1c, % | 6.4±1.0 | 5.9±0.6 | 6.3±0.9 | 6.0±0.6 | 6.0±0.8 | 5.9±0.7 | <0.001 |
| Serum creatinin, mg/dL | 1.59±2.12 | 1.03±1.36 | 1.65±1.78 | 1.78±2.29 | 1.06±0.50 | 1.31±1.70 | <0.001 |
| Serum sodium, mEq/L | 140.5±2.6 | 141.3±2.1 | 140.0±3.7 | 140.0±2.9 | 140.1±3.0 | 140.6±2.6 | <0.001 |
| NT-proBNP, pg/mL | 2066±8730 | 581±1671 | 7170±23314 | 5349±18189 | 923±1389 | 2116±11782 | <0.001 |
| CTR (chest X-ray), % | 48.1±5.4 | 47.4±5.4 | 54.4±7.0 | 51.7±6.6 | 50.9±6.4 | 50.1±6.9 | <0.001 |
| LVEF, % | 60.1±12.9 | 65.0±8.6 | 46.3±18.0 | 62.1±12.6 | 67.5±8.9 | 61.1±14.0 | <0.001 |
| Female | Coronary artery disease | Arrhythmia | Heart failure | Valvular heart disease | Pulmonary hypertension | Others | P for trend |
| n (%) | 437 (19.5) | 552 (24.6) | 544 (24.3) | 177 (7.9) | 257 (11.5) | 275 (12.3) |  |
| Age, year | 71.0±12.1 | 64.3±14.7 | 73.0±13.0 | 76.8±11.6 | 60.5±16.2 | 59.9±16.5 | <0.001 |
| SpO2 at awakening, % | 94.1±2.6 | 94.4±1.8 | 93.9±2.5 | 94.2±1.8 | 92.3±3.6 | 94.2±2.7 | <0.001 |
| Systolic blood pressure, mmHg | 125.2±18.4 | 123.3±17.4 | 118.2±20.5 | 125.3±19.9 | 110.2±16.1 | 119.1±18.9 | <0.001 |
| Diastolic blood pressure, mmHg | 68.4±11.4 | 72.6±11.7 | 67.0±13.5 | 68.8±11.9 | 66.6±11.7 | 70.4±12.9 | <0.001 |
| Pulse pressure, mmHg | 56.7±15.6 | 50.7±15.5 | 51.2±17.7 | 56.5±16.7 | 43.7±11.7 | 48.6±14.5 | <0.001 |
| Pulse rate, bpm | 70.5±11.2 | 71.8±17.1 | 75.9±15.3 | 74.7±10.9 | 81.2±13.5 | 75.7±13.9 | <0.001 |
| BMI, kg/m2 | 23.7±4.3 | 23.1±3.6 | 22.7±4.3 | 22.1±3.5 | 22.2±4.6 | 22.4±4.2 | <0.001 |
| WBC, /μL | 5979±1997 | 5563±1543 | 5992±2691 | 5731±1730 | 5264±2014 | 5709±2126 | 0.021 |
| Hb, g/dL | 12.2±1.5 | 13.0±1.3 | 11.9±1.9 | 12.0±1.6 | 12.2±1.9 | 12.7±1.6 | <0.001 |
| HbA1c, % | 6.4±0.9 | 5.9±0.6 | 6.1±0.8 | 5.9±0.8 | 5.8±0.6 | 5.8±0.6 | <0.001 |
| Serum creatinin, mg/dL | 1.17±1.53 | 0.77±0.90 | 1.24±1.77 | 0.80±0.60 | 0.78±0.85 | 0.83±1.08 | <0.001 |
| Serum sodium, mEq/L | 141.1±2.9 | 141.5±2.1 | 140.4±3.4 | 141.1±2.8 | 141.0±2.6 | 140.8±2.6 | <0.001 |
| NT-proBNP, pg/mL | 3626±20097 | 677±2453 | 6202±16230 | 1834±4270 | 850±1745 | 1369±6066 | <0.001 |
| CTR (chest X-ray), % | 51.7±5.7 | 49.9±6.2 | 58.4±7.6 | 54.2±6.4 | 53.4±7.3 | 52.4±7.5 | <0.001 |
| LVEF, % | 63.6±12.4 | 67.5±8.1 | 54.8±18.7 | 67.1±10.0 | 69.4±8.4 | 65.2±12.5 | <0.001 |

Further analysis of CVD prevalence within SDB categories revealed that, in all patients, coronary artery disease was the most prevalent, followed by arrhythmia and heart failure in mild SDB cases. In moderate and severe SDB categories, coronary artery disease remained predominant, with heart failure and arrhythmia following in prevalence (**Table 4**, **Figure 2**). For males, the prevalence pattern of underlying CVDs mirrored the overall findings. However, for females, arrhythmia and heart failure were the most prevalent, succeeded by coronary artery disease, pulmonary hypertension, and valvular heart disease. In moderate to severe SDB, heart failure took precedence, followed by coronary artery disease, arrhythmia, valvular heart disease, and pulmonary hypertension (**Table 4**, **Figure 2**).

**Figure 2.**
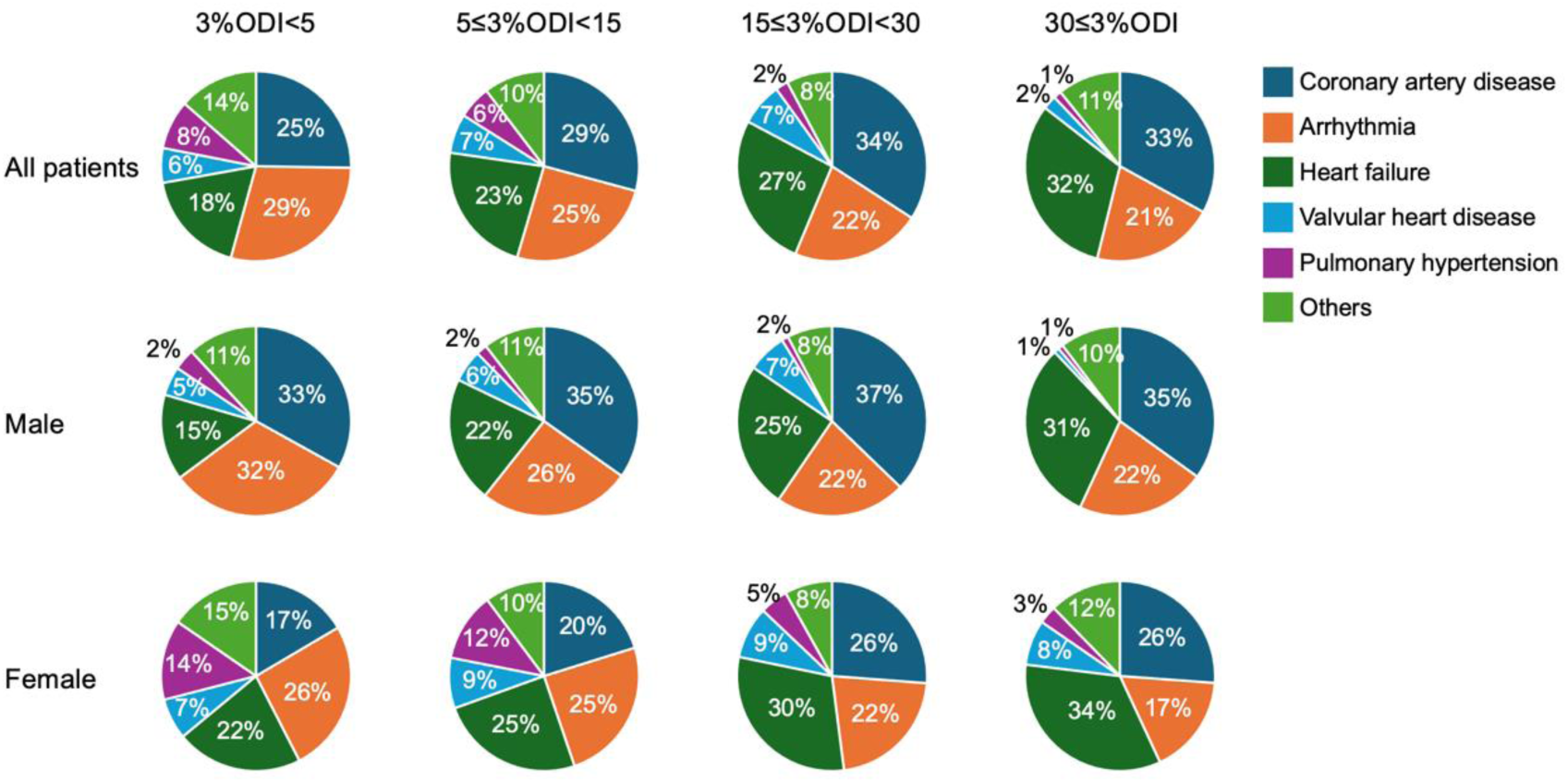
Underlying cardiovascular diseases in each SDB severity group. Top; all patients, middle; male, bottom; female.

**Table 4.** Severity of sleep-disordered breathing in each cardiovascular diseases.

|  | Total | Coronary artery disease | Arrhythmia | Heart failure | Valvular heart disease | Pulmonary hypertension | Others | P for trend |
| --- | --- | --- | --- | --- | --- | --- | --- | --- |
| 3%ODI<5 | 2067 | 522 | 600 | 370 | 122 | 173 | 280 | <0.001 |
| 5≤3%ODI<15 | 2426 | 708 | 616 | 551 | 162 | 137 | 252 |  |
| 15≤3%ODI<30 | 975 | 333 | 216 | 258 | 71 | 21 | 76 |  |
| 30≤3%ODI | 297 | 98 | 62 | 94 | 7 | 4 | 32 |  |
| Male |  | Coronary artery disease | Arrhythmia | Heart failure | Valvular heart disease | Pulmonary hypertension | Others |  |
| 3%ODI<5 | 1092 | 361 | 347 | 160 | 54 | 40 | 130 | <0.001 |
| 5≤3%ODI<15 | 1495 | 520 | 387 | 321 | 82 | 28 | 157 |  |
| 15≤3%ODI<30 | 704 | 262 | 157 | 176 | 47 | 8 | 54 |  |
| 30≤3%ODI | 232 | 81 | 51 | 72 | 2 | 2 | 24 |  |
| Female |  | Coronary artery disease | Arrhythmia | Heart failure | Valvular heart disease | Pulmonary hypertension | Others |  |
| 3%ODI<5 | 975 | 161 | 253 | 210 | 68 | 133 | 150 | <0.001 |
| 5≤3%ODI<15 | 931 | 188 | 229 | 230 | 80 | 109 | 95 |  |
| 15≤3%ODI<30 | 271 | 71 | 59 | 82 | 24 | 13 | 22 |  |
| 30≤3%ODI | 65 | 17 | 11 | 22 | 5 | 2 | 8 |  |

The data are presented as the number.

Lastly, the prevalence of SDB within each CVD category was assessed (**Table 4**, **Figure 3**). In coronary artery disease, 30% of patients exhibited no SDB, nearly half had mild SDB, 20% moderate SDB, and 5-6% severe SDB). For arrhythmia, a combination of no and mild SDB was observed in 80% of cases. In heart failure and valvular heart disease, no SDB was observed in 20-30% of cases, nearly half presented with mild SDB, 20% with moderate SDB, and less than 10% with severe SDB. In pulmonary hypertension, over half of the patients exhibited no SDB, with 40% displaying mild SDB (**Table 4**, **Figure 3**).

**Figure 3.**
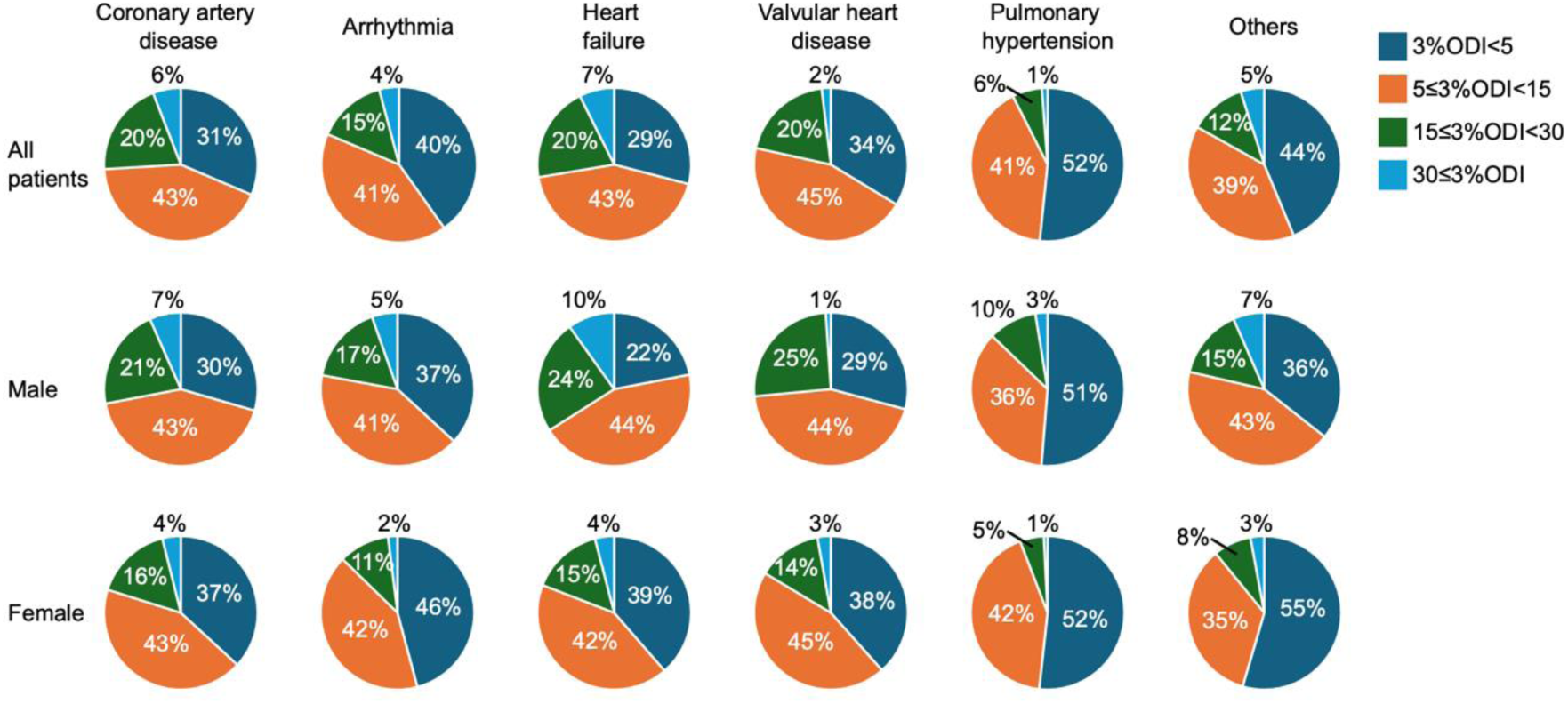
Severity of SDB in each underlying cardiovascular disease. Top; all patients, middle; male, bottom; female.

### Heatmaps of Oxygen Saturation Levels and Body Mass Index in Relation to the Severity of Sleep-Disordered Breathing

As shown in **Figure 4**, the heatmaps illustrate the relationship between SpO2 upon awaking and BMI, with the color gradient indicating the percentage of severe SDB cases (fewer severe cases in green, more severe cases in red). It visually demonstrates that lower awake SpO2 and higher BMI are associated with an increased percentage of severe SDB cases (**Figure 4**).

**Figure 4.**
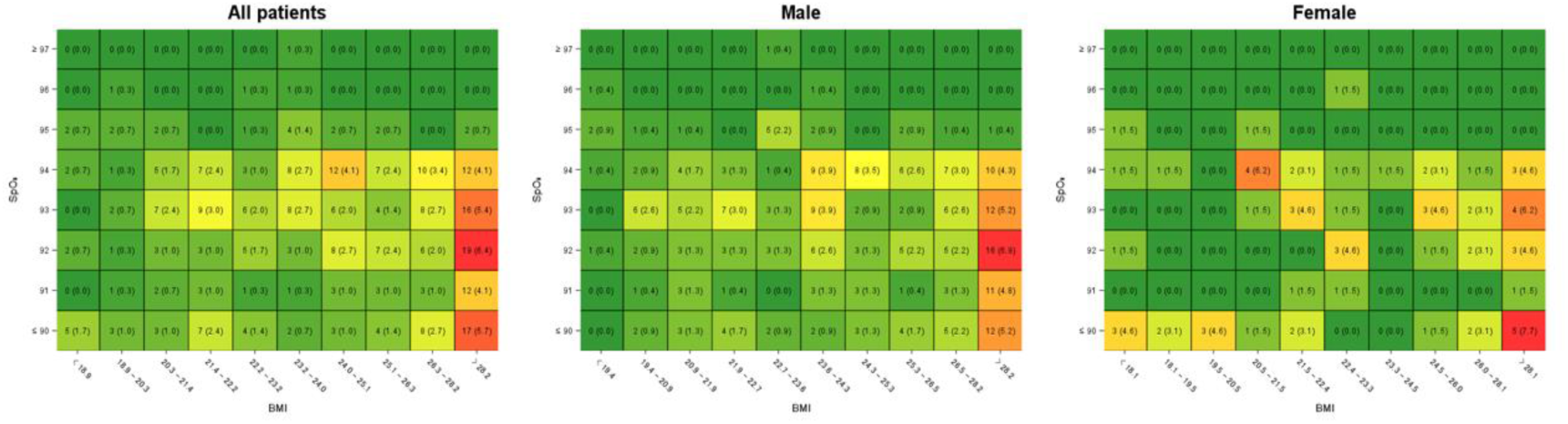
Heatmaps of Oxygen Saturation Levels and Body Mass Index in Relation to the Severity of Sleep-Disordered Breathing. The x-axis represents body mass index (BMI), categorized into various ranges, while the y-axis represents SpO_2_ levels upon awakening. The color intensity indicates the percentage of severe sleep disordered breathing (SDB), with green representing fewer severe SDB cases and red indicating more severe cases. As the color transitions from green to red, the percentage of severe SDB increases, visually emphasizing that patients with elevated BMI and reduced SpO_2_ are at significant risk for severe SDB.

## Discussion

To our knowledge, this is the first study to juxtapose the severities of SDB across a spectrum of CVDs utilizing a singular database. Notably, we discovered that patients with more severe SDB exhibited lower SpO_2_ upon awakening, specifically within the context of CVDs. The combination of higher BMI and lower SpO_2_ levels upon awakening can predict the presence of severe SDB. The other primary findings of our investigation can be encapsulated as follows: (1) Mirroring the general population, being male and of advanced age emerged as risk factors for SDB among CVD patients. (2) The analysis was stratified by sex and age to furnish a granular understanding of SDB susceptibility. (3) Across both sex and age brackets, a correlation was established between the severity of SDB and both hypoxia saturation at awakening and BMI. (4) Elevated HbA1c levels were associated with SDB in females in their 70s and those aged 80 and above. (5) An increased cardiothoracic ratio, as assessed via chest radiography, was linked with heightened SDB severity among all male cohorts and females over 80 years of age. Furthermore, reduced LVEF, as determined by echocardiography, was associated with SDB across all male groups and females in their 70s. The overarching theme underscores a strong correlation between traditionally adverse clinical parameters and the presence of SDB.

### Oxygen Saturation upon Awakening: A Potential Predictor of SDB Severity

In our analysis, the average SpO_2_ at awakening was 93.2 ± 2.1%. Sex-specific analysis revealed that males had an average SpO_2_ of 93.4 ± 2.0% and females 92.7 ± 2.3% upon awakening, with both showing moderate to severe SDB. Our findings indicate that lower daytime SpO_2_ is associated with more severe SDB, a trend observable regardless of sex or age. Thus, SpO_2_ upon awakening may serve as a reliable marker for SDB, especially in control subjects with CVDs undergoing hospital-based diagnostic and therapeutic procedures.

### Hypertension, Obesity, and Sleep-Disordered Breathing

The relationship between systolic and diastolic blood pressures, as well as pulse pressure, and SDB appeared to be weak in our study.^7–9^ Although there is a well-documented link between hypertension and SDB in previous reports, such an association was not evident in our cohort, probably because our participants with hypertension were already under treatment with antihypertensive medications. Consequently, the therapeutic control of hypertension might obscure the detection of any direct association between blood pressure and SDB.

In contrast, a significant correlation between pulse rate and SDB was observed; however, it was limited to the overall group under 59 years of age and specifically among males in this age group. Our findings suggest that an elevated heart rate is associated with the severity of SDB in the younger age groups. It is important to note that the pulse rate measurements referenced in this study were taken upon awakening and at rest, as previously reported.^10^

Consistent with these findings,^11,12^ our study demonstrated that higher BMI levels were associated with greater severity of SDB across all demographic groups. This relationship was evident regardless of sex or age, underscoring the strong influence of obesity on SDB severity. As shown in **Figure 4**, the combination of BMI and daytime SpO_2_ may predict the severe SDB.

### Blood Test and Sleep-Disordered Breathing

In our analysis, white blood cell counts showed correlations with SDB across all age groups, except males aged over 80. Among females, significant correlations were observed only in those under 59 years. Conversely, when age was not considered in the analysis, a correlation was evident. While previous studies have suggested a link between WBC count and SDB, our findings indicate that white blood cell count may not serve as a reliable indicator when categorized by age group. Given the variability of this association across different age brackets,^13^ a nuanced analysis that takes into account both sex and age is recommended for a more accurate assessment.

The relationship between hemoglobin levels and SDB in our study was weak. Although SDB can lead to polycythemia vera due to hypoxia-induced erythrocytosis,^14–16^ hemoglobin levels may not be a dependable marker for SDB, particularly in patients with CVDs who often exhibit anemia as a consequence of their underlying condition.

Regarding HbA1c, a strong correlation with SDB was identified in most groups, except for females over 70 and males over 80. The association between diabetes mellitus and SDB, as demonstrated in numerous studies,^17–19^ was also reflected in our study of patients with CVDs.

The correlation between serum creatinine levels and SDB was confirmed in the overall analysis;^20,21^ however, when segmented by age group, the correlation persisted only in males under 59 years and females under 69 years. This discrepancy might be attributed to the inclusion of dialysis patients in our study, many of whom had impaired renal function secondary to CVDs. Consequently, serum creatinine may not be a reliable marker in elderly patients with CVDs.

Concerning NT-proBNP, the overall analysis showed a correlation, particularly in females under 59 and males in their 60s. Although continuous positive airway pressure therapy for sleep apnea has been reported to reduce NT-proBNP levels in some studies, others have found no association.^22,23^ The variability observed in our study, which included patients with CVDs who generally have elevated baseline NT-proBNP levels, suggests that NT-proBNP may not be a definitive indicator in the presence of CVDs.

### Cardiac Function and Sleep-Disordered Breathing

The cardiothoracic ratio, as measured by chest X-ray, demonstrated significant correlations with SDB across all male age groups and in all female age groups, with the exception of those aged 60-79 years. Notably, this correlation persisted in both males and females aged over 80 years, a demographic in which few parameters were found to correlate with SDB. Several studies have identified a link between obstructive sleep apnea and cardiac hypertrophy,^24,25^ suggesting that the cardiothoracic ratio could serve as a reliable indicator of SDB.

Furthermore, a relationship between reduced LVEF and the severity of sleep apnea has been documented.^26,27^ In our study, a decreased LVEF, as determined by echocardiography, was associated with SDB in all male age groups. Among females, this association was observed specifically in those in their 70s. Previous research has shown a correlation between SDB and decreased LVEF in healthy individuals, indicating that LVEF could be a pertinent indicator of SDB severity, particularly in males with CVDs.

### Perspective of Disease-Specific Profiles

Our analysis revealed that factors such as sex, age, SpO_2_ upon awakening, BMI, and HbA1c levels are associated with the severity of SDB. However, when examining the impact of specific CVDs, heart failure and coronary artery disease emerged as conditions with a higher prevalence and severity of SDB compared to others. Specifically, valvular heart disease, typically linked with older age, and pulmonary hypertension, associated with lower SpO_2_ levels upon awakening, exhibited lower incidences and severities of SDB. These findings suggest that coronary artery disease and heart failure might be particularly susceptible to the development and exacerbation of SDB.

### Limitations

This study has several limitations. First, the focus is exclusively on patients with CVDs, rather than the general population. Consequently, many findings reported for the general population may not be directly applicable or relevant to our study. Second, the findings are pertinent primarily to hospitalized patients undergoing evaluation and treatment for CVDs, limiting the applicability of our results to this specific patient group. Moreover, the study groups included a mix of conditions -coronary artery disease, arrhythmias, heart failure, and pulmonary hypertension-without a detailed categorization or background analysis. Future research should aim to dissect and validate findings by individually analyzing each disease type for a more precise understanding. Third, SDB encompasses two distinct conditions: OSA and CSA, each with unique mechanisms. Our methodology, relying on PULSOX®-Me, was not able to differentiate between these types, a significant limitation given the importance of distinguishing between OSA and CSA for accurate diagnosis and management.

## Conclusions

The current study elucidates the clinical profiles of CVD patients exhibiting more severe forms of SDB. Our disease-specific analysis revealed that coronary artery disease and heart failure are associated with a higher prevalence and a greater risk of moderate to severe SDB compared to valvular heart diseases, typically in older patients. Notably, low oxygen saturation during daytime and elevated BMI were indicative of SDB across all age categories for both sexes.

## Data Availability

The data presented in this study are available on request from the corresponding author. The data are not publicly available due to privacy restrictions.

## Acknowledgments

None.

## Data availability statement

The data underlying this article will be shared with others for the purpose of reproducing results or replicating procedures upon reasonable request to the corresponding author, subject to institutional and ethics committee approval.

## Authors’ Contributions

Koutatsu Shimozono: Conceptualization, Methodology, Formal analysis, Data Curation, and Writing-Original draft preparation.

Hisashi Adachi: Statistics.

Shoichiro Nohara: Methodology, Investigation, Writing-Review and editing.

Tatsuhiro Shibata: Methodology, Data Curation, and Formal analysis.

Yoichi Sugiyama: Investigation.

Yoshihiro Fukumoto: Project administration, Conceptualization, Writing-Review and editing.

All authors have read and approved the final article.

## Abbreviations list

AHI: apnea-hypopnea index
ANOVA: analysis of variance
BMI: body mass index
CPAP: continuous positive airway pressure
CVDs: cardiovascular diseases
HbA1c: hemoglobin A1c
LVEF: left ventricular ejection fraction
NT-proBNP: N-terminal pro B-type natriuretic peptide
ODI: oxygen desaturation index
OSA: obstructive sleep apnea
SBP: systolic blood pressure
SD: standard deviation
SDB: sleep-disordered breathing.
SpO_2_: transcutaneous arterial blood oxygen saturation

